# Utilization of health care services before and after media attention about fatal side effects of the AstraZeneca vaccine: a nation-wide register-based event study

**DOI:** 10.1101/2021.07.09.21260250

**Authors:** Vilde Bergstad Larsen, Mari Grøsland, Kjetil Telle, Karin Magnusson

**Author notes:** These authors contributed equally to the study (Vilde Bergstad Larsen, Mari Grøsland). **Corresponding author, contact information:** Vilde Bergstad Larsen, Cluster for Health Services Research, Norwegian Institute of Public Health, Postboks 222, Skøyen, N-0213 Oslo. Visiting address: Sandakerveien 24c, Building D, 0473 Oslo.

## Abstract

**Background:** Survey studies have found that vaccinated persons tend to report more side effects after being given information about side effects rather than benefits. However, the impact of high media attention about vaccine-related side effects on the utilization of health care is unknown. We aimed to assess whether utilization of health care services for newly vaccinated health care workers changed after media attention about fatal side effects of the AstraZeneca vaccine on March 11^th^, 2021, and whether changes differed by age, sex, or occupation.

**Methods:** We utilized individual-level data on health care use, vaccination, employment, and demographics available in the Norwegian emergency preparedness register Beredt C19. In all 99,899 health care workers in Norway who were vaccinated with AstraZeneca between February 11^th^ and March 11^th^, we used an event-study design with a matched comparison group to compare the change in primary and inpatient specialist care use from 14 days before to 14 days after the information shock on March 11^th^, 2021.

**Results:** Primary health care use increased with 8.2 daily consultations per 1,000 health care workers (95% CI 7.51 to 8.89) the week following March 11^th^ for those vaccinated with AstraZeneca (n = 99,899), compared with no increase for the unvaccinated comparison group (n = 186,885). Utilization of inpatient care also increased with 0.8 daily hospitalizations per 1,000 health care workers (95% CI 0.37 to 1.23) in week two after March 11^th^. The sharpest increase in daily primary health care use in the first week after March 11^th^ was found for women aged 18-44 (10.6 consultations per 1,000, 95% CI 9.52 to 11.68) and for cleaners working in the health care sector (9.8 consultations per 1,000, 95% CI 3.41 to 16.19).

**Conclusions:** Health care use was higher after the media reports of a few cases of fatal or severe side effects of the AstraZeneca vaccine. Our results suggest that the reports did not only lead vaccinated individuals to contact primary health care more, but also that physicians referred and treated more cases to specialist care after the new information.

## Background

Norwegian health authorities suspended the use of the Oxford-AstraZeneca (Vaxevria, hereafter referred to as AstraZeneca) vaccine on March 11^th^, 2021, after sudden reports of severe and sometimes fatal thromboembolic events following vaccination were announced earlier the same day.^1^ In these announcements, it became clear that the European Medicines Agency (EMA) had received reports of 30 similar events among the five million individuals in the European Economic Area who had received the AstraZeneca vaccine.^2^ While it was still not known whether the thromboembolic events could be linked to the vaccine, the severe cases and fatalities among otherwise healthy individuals received much media attention in the days that followed.

In Norway, AstraZeneca was administered to more than 100,000 health care workers (HCWs, including cleaners in the health care sector) under the age of 65 until March 11^th^, 2021. Multiple cases of thromboembolism and four deaths among vaccinated HCWs, predominately young women, were discovered after March 11^th^, which eventually resulted in the permanent removal of AstraZeneca from the Norwegian COVID-19 vaccination program on May 10^th^, 2021.^3^ Along the fatal events, recent survey studies based on self-reporting, conducted after March 11^th^, have also reported minor bleeding episodes such as skin and nose bleedings following vaccination with AstraZeneca.^4^

The unanticipated reporting of serious side effects on March 11^th^ provided an opportunity to better understand how an *information shock* (high media attention, particularly about the fatalities, and official encouragements to seek medical attention if experiencing a range of symptoms) affects the utilization of health care services. More specifically, it allowed us to study how health-literate HCWs responded to unexpected reports of low-probability severe and sometimes fatal outcomes.

Assessing how vaccinated individuals react to sudden media coverage of vaccine-related side effects is important to ensure efficient health care services in similar situations in the future. To this end, we aimed to compare the utilization of health care services following AstraZeneca vaccination before and after the information shock on March 11^th^. Furthermore, we explored variation in responses to the information shock across age, sex, and health care professions.

## Methods

Using a pre-post study design with a comparison group and multiple time periods, we utilized data from the BEREDT C19 registry, which is a newly developed emergency preparedness register aiming to provide rapid knowledge about the COVID-19 pandemic, including how measures enforced to limit the spread of the virus affect the population’s health, use of health care services and health-related behaviors.^5^ From within BEREDT C19, we utilized nation-wide individual-level data originating from the following registries: The National Population Register; Norway Control and Payment of Health Reimbursement (KPR, visits to general practitioners and emergency wards); Norwegian Patient register (NPR, outpatient and inpatient hospital care); the Employer- and Employee Register (all employment contracts, used to define sample of HCW); the Norwegian Surveillance System for Communicable Diseases (MSIS, all polymerase chain reaction tests for SARS-CoV-2), and the Norwegian Immunization Registry (SYSVAK, vaccination date and vaccine manufacturer). Individuals could be linked across data sources and over time using an encrypted version of the unique personal identification number provided every resident of Norway at birth or upon first immigration. Additionally, we included data from the media monitoring company Retriever on the daily number of published media reports in Norway that included the words “AstraZeneca” and “side effect”.

### Study sample

Our study population included all Norwegian residents, aged 18-65, who had an employment contract as HCW on February 11^th^, 2021, and who had not received any COVID-19 vaccine or tested positive for COVID-19 by February 11^th^, 2021 (Fig. A1).

We identified HCW following Molvik et al. by using the ISCO-08 4-digit occupation codes, in combination with standard industrial classifications from the Employer- and Employee-register (see appendix of Molvik et al. for the exact definition).^6^ We divided our population into two mutually exclusive groups:

1. HCWs vaccinated with AstraZeneca from February 11^th^ to March 11^th^, 2021 (treatment group)
2. A comparison group of propensity-score matched HCW, where we matched each HCW in our treatment group to two (nearest-neighbor) HCWs who had not received the AstraZeneca vaccine, based on their estimated probability of being vaccinated with AstraZeneca via a logistic regression model (see Appendix A for details).

The HCWs in the comparison group were assigned the vaccination date of their vaccinated match. Because the rare conditions that prompted the pausing of AstraZeneca were reported to have occurred shortly after vaccination, we observed HCWs for 14 days after their (hypothetical) vaccination date.^7, 8^ HCWs in both groups who tested positive for COVID-19, died, or emigrated were observed only until the date of positive test, death, or emigration. Any person-time after the event was excluded from further analysis. When these events occurred prior to the (hypothetical) vaccination date, the HCWs were excluded entirely, as they could not contribute to the 14-day observation period. Similarly, HCWs in the comparison group who received a different vaccine than AstraZeneca, were observed only until the date of actual vaccination. Hence, the comparison group consisted solely of HCWs who were unvaccinated by the hypothetical vaccination date.

To evaluate the impact of the information shock on March 11^th^ on health care use, we compared the daily health care use of HCWs in the treatment and comparison groups, from 14 days before March 11^th^, (i.e., February 25^th^), to 14 days after March 11^th^, (i.e., March 25^th^). This means that outcomes for persons who were vaccinated on February 11^th^ were observed on February 25^th^ only, persons who were vaccinated on February 12^th^ were observed February 25^th^ and 26^th^, and so on. Similarly, persons who were vaccinated on March 10^th^ contributed with observations for 14 days from March 11^th^ to March 24^th^.

### Outcomes

We studied all health care contacts with primary and specialist care combined, as well as separately:

1. Primary care included consultations at the general practitioner and emergency ward as well as outpatient hospital contacts.
2. Specialist care included inpatient hospital contacts (overnight hospitalization).

The categorical outcome variables were measured daily and set to one if the HCW had a health care contact the given day (otherwise zero).

### Statistical analyses

To examine the impact of the information shock on health care use, we compared HCWs who were vaccinated with AstraZeneca (treatment group) to the HCWs in the comparison group. First, we calculated and plotted the daily health care use (primary and specialist care) per 1,000 HCWs from February 25^th^ to March 25^th^, i.e., from two weeks before to two weeks after the information shock on March 11^th^. This was calculated by dividing the daily number of HCWs with a health care contact by the number of HCWs with or without a health care contact the given day, and then multiplied by 1,000. We did this separately for the vaccinated HCWs and the HCWs in the comparison group. To illustrate that the information about fatal side effects of the AstraZeneca vaccine was indeed new on March 11^th^, we also plotted the daily number of published media reports in Norway that mentioned both “AstraZeneca” and “side effects”. Second, we studied the impact of the information shock on primary care use and specialist care use in separate analyses using an event study design operationalized as a difference-in-differences (DiD) approach with separate estimates for each week (or day, see below) after March 11^th^.^9^ This allowed us to assess the change in health care use each week from before March 11^th^ to after March 11^th^ for those who had received the AstraZeneca vaccine relative to those in the comparison group. With this method we could effectively adjust for the substantial calendar week-day variation in health care utilization.

DiD analyses evaluate the effect of an event by comparing the change in the outcome for the affected group before and after the event, to the change over the same time span in a group not affected by the event.^9-12^ In this study, we compared the daily health care use per 1,000 HCWs in the 14-day period before and two 7-day periods after March 11^th^ for the HCWs who were vaccinated (difference 1), to the daily difference in health care use per 1,000 HCWs in the 14-day period before and two 7-day periods after the March 11^th^ for the HCWs in the comparison group (difference 2). The DiD estimate is the difference between these two differences, estimated using linear regression with robust standard errors and presented as an absolute difference in daily health care use per 1,000 HCWs. If there is no increase in health care use from before March 11^th^ to after March 11^th^, the DiD estimate would be zero.

We thus extended the traditional DiD method with one period before and one period after the event to two separate estimates for each 7-day period after March 11^th^, comparing to the health care use in the 14-day period before March 11^th^.^9-14^ In addition to this model with separate estimates for the first and second week after March 11^th^, we also estimated more detailed models with separate estimates for each calendar day before and after March 11^th^, comparing to the health care use on March 11^th^. The results from such models with estimates for each calendar day were presented in plots. In addition to the presentation of results as absolute differences per 1,000 HCWs, we also presented relative differences (i.e. in percent) by dividing the absolute estimate for each of the post periods by the health care use of the comparison group in the reference pre period (multiplied by 100). Our outcome variables were either measured jointly as daily health care use during the first and second week after March 11^th^ or by calendar day. To handle that a HCW can be present several times in the data, robust standard errors were clustered on the individual level.

To investigate group differences, we split our main sample into four mutually exclusive groups; women aged 18-44, women aged 45-65, men aged 18-44 and men aged 45-65 and ran the same regressions as above on these groups. We also conducted the same analysis on HCWs in five mutually exclusive occupational groups (physicians, health professionals, health associate professionals, personal care workers, and cleaners) based on the first digit in the ISCO-08 classification (except for physicians).^15^ All analyses were run in R v.3.6.2.

### Ethics

The establishment of an emergency preparedness register forms part of the legally mandated responsibilities of The Norwegian Institute of Public Health (NIPH) during epidemics. Institutional board review was conducted, and the Ethics Committee of South-East Norway confirmed (June 4th, 2020, #153204) that external ethical board review was not required.

## Results

Among 357,685 persons registered with an employment contract as HCW on February 11^th^, 2021, we studied the treatment group of 99,883 persons who had received the AstraZeneca vaccine in the time period from February 11^th^ to March 11^th^ and a comparison group of their 186,387 unvaccinated matches (Table 1). As intended, there were only small differences on variables predicting AstraZeneca vaccination between the treatment and comparison group. Both groups consisted of a majority of younger, Norwegian-born women working as personal care assistants or health professionals (Table 1).

**Table 1.** Descriptive statistics

|  | **AstraZeneca** | **Comparison group** |
| --- | --- | --- |
| N | 99,883 | 186,387 |
| Person-days | 1,356,733 | 2,520,648 |
| Mean (SD) age | 42.3 (12.9) | 41.9 (12.9) |
| Women, N (%) | 82,457 (82.6) | 156,602 (84) |
| Norwegian born, N (%) | 82,631 (82.7) | 148,197 (79.5) |
| **Occupational groups** |  |  |
| Physicians, N (%) | 6,226 (6.2) | 11,882 (6.4) |
| Health professionals, N (%) | 33,522 (33.6) | 59,676 (32.0) |
| Health associate professionals, N (%) | 6,110 (6.1) | 9,794 (5.3) |
| Personal care workers, N (%) | 52,989 (53.1) | 102,995 (55.3) |
| Cleaners, N (%) | 1,036 (1.0) | 2,040 (1.1) |
| **Daily health care use per 1,000 HCWs** |  |  |
| Two weeks prior to March 11<sup>th</sup> |  |  |
| All health care | 10.5 | 13.4 |
| Primary care | 10.2 | 12.4 |
| Inpatient specialist care | 0.3 | 1.2 |
| First week after March 11<sup>th</sup> |  |  |
| All health care | 19 | 13.7 |
| Primary care | 18.5 | 12.5 |
| Inpatient specialist care | 0.7 | 1.3 |
| Second week after March 11<sup>th</sup> |  |  |
| All health care | 18.5 | 12.9 |
| Primary care | 17.9 | 12 |
| Inpatient specialist care | 0.9 | 1.0 |

*SD: Standard deviation*

The daily health care utilization in the period prior to the information shock was 10.5 consultations per 1,000 persons for the treatment group vaccinated with AstraZeneca, while the corresponding numbers for March 12^th^-18^th^ and March 19^th^-25^th^ were 19.0 and 18.5 consultations per 1,000 persons (Table 1).

The information shock occurred on March 11^th^, with more than 125 news articles covering AstraZeneca-related side effects, in stark contrast to the very small number of media reports that mentioned “AstraZeneca” and “side effect” prior to this date (Fig. 1, secondary axis). The following day, we observed an increase in daily health care use for those vaccinated with AstraZeneca compared to the HCWs in our comparison group which continued in the days that followed (Fig. 1, primary axis). While the comparison group had slightly higher average health care use prior to March 11^th^ than the treatment group, the comparison group had no increase in health care utilization from before to after March 11^th^ (Table 1, Fig. 1), which indicates that the increase for individuals vaccinated with AstraZeneca was not due to any other acute events occurring around the same date.

**Figure 1.**
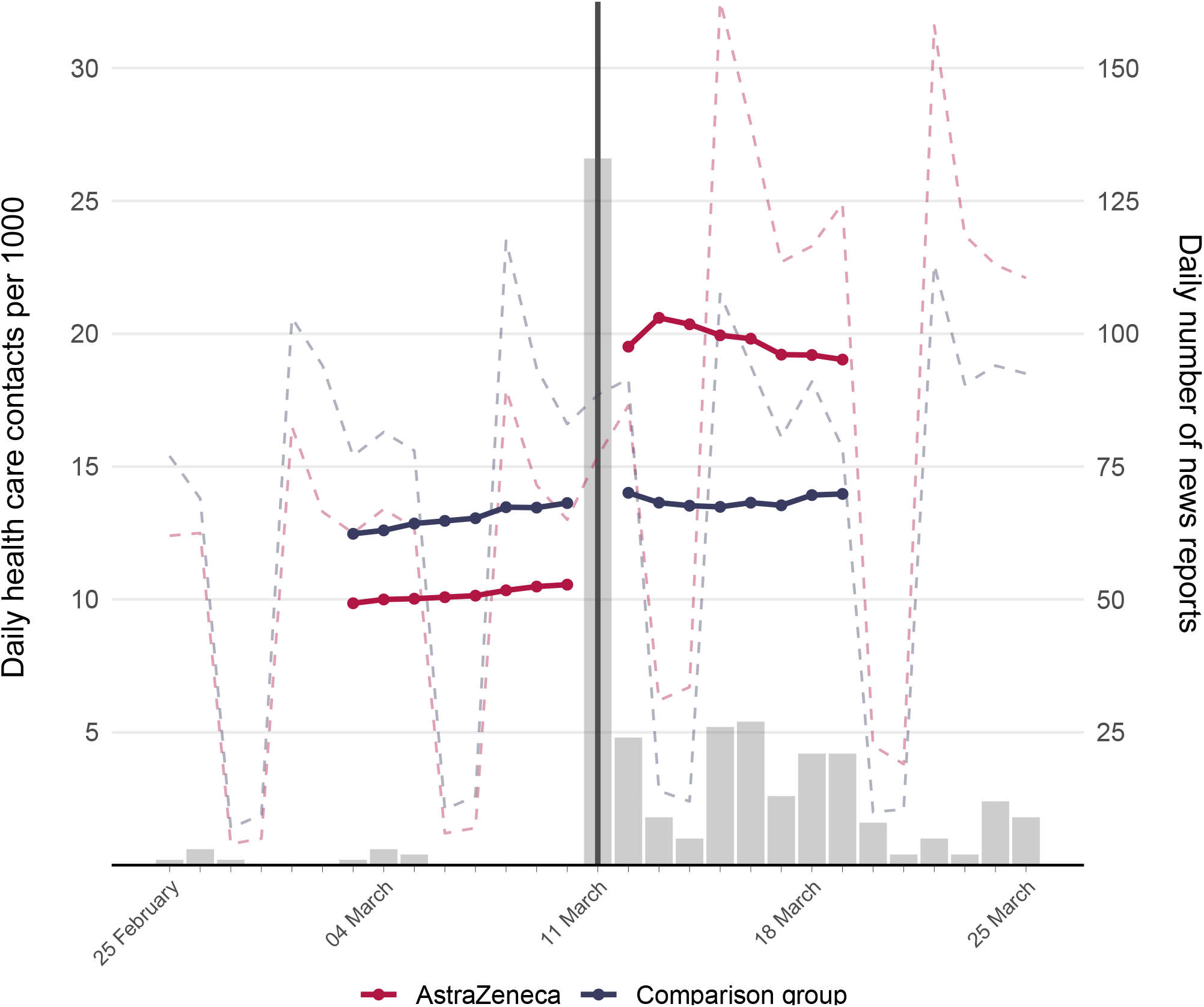
Average daily health care utilization and the information shock on March 11^th^ Daily (dashed lines) and 7-day rolling average (solid lines) of daily health care use after vaccination. The 7-day rolling average was calculated separately before and after March 11^th^ to visualize the change in health care use after the media attention (bars, secondary axis). For days prior to and including March 11^th^, the 7-day rolling average equals the mean of health care use per 1,000 HWCs the given day and the six preceding days, while for days after March 11^th^, it equals the mean of health care use per 1,000 HCWs the given day and the six subsequent days. Bars indicate daily number of news reports mentioning “AstraZeneca” and “side effect”.

### Impact of the information shock on health care use after AstraZeneca vaccination

Primary health care use for HCWs vaccinated with AstraZeneca increased with 8.2 daily consultations per 1,000 HCWs in the first week after March 11^th^ when compared to HCWs in the comparison group (Table 2). This corresponded to a 66 % increase compared to levels prior to March 11^th^ (Table 2). A similar increase of 8.1 per 1,000 HCWs (65 %) was observed in the second week after March 11^th^ (Table 2). Estimates of the daily difference between use of primary care for those vaccinated vs. the comparison group rose from the first day after March 11^th^, peaking after around 4-5 days (Fig. 2). We also observed an increase of 0.2 per 1,000 HCWs (20 %) in inpatient specialist care visits the first week, and an increase of 0.8 per 1,000 HCWs (66 %) in the second week after March 11^th^ (Table 2, Fig. 2).

**Table 2.** Impact of the information shock on health care use

| Period after<br>March 11<sup>th</sup> | Primary Care |  |  |  | Inpatient specialist care |  |  |  |
| --- | --- | --- | --- | --- | --- | --- | --- | --- |
| | $\beta$ | St. err. | % Relative diff<br>($\beta$) | Relative diff<br>(St. err.) | $\beta$ | St. err. | % Relative diff<br>($\beta$) | Relative diff<br>(St. err.) |
| First week | 8.2*** | 0.35 | 66 | 2.8 | 0.2 | 0.15 | 20 | 12.9 |
| Second week | 8.1*** | 0.53 | 65 | 4.3 | 0.8*** | 0.22 | 66 | 18.6 |

*Notes:* Differences-in-differences estimates for the change in daily health care use per 1,000 persons for different health care services before and after March 11<sup>th</sup> for HCWs vaccinated the past 14 days. In primary care reimbursement codes 2ad, 2ak, 2ae is used to identify consultations at general practitioner or emergency ward. All models control for age and sex. Standard errors (St. err.) are clustered on individuals. The pre-period (health care utilization after vaccination the two weeks prior to March 11<sup>th</sup>) is reference period in all regressions. In addition to the presentation of results as absolute differences per 1,000 persons, relative differences (i.e., in percent) are also presented, calculated by dividing the absolute estimate (and corresponding standard error) for each of the post periods by the health care use rate of the comparison group in the pre period (and multiplying with 100).
Significance levels: \* <0.1; \*\* <0.05; \*\*\* <0.01

**Figure 2.**
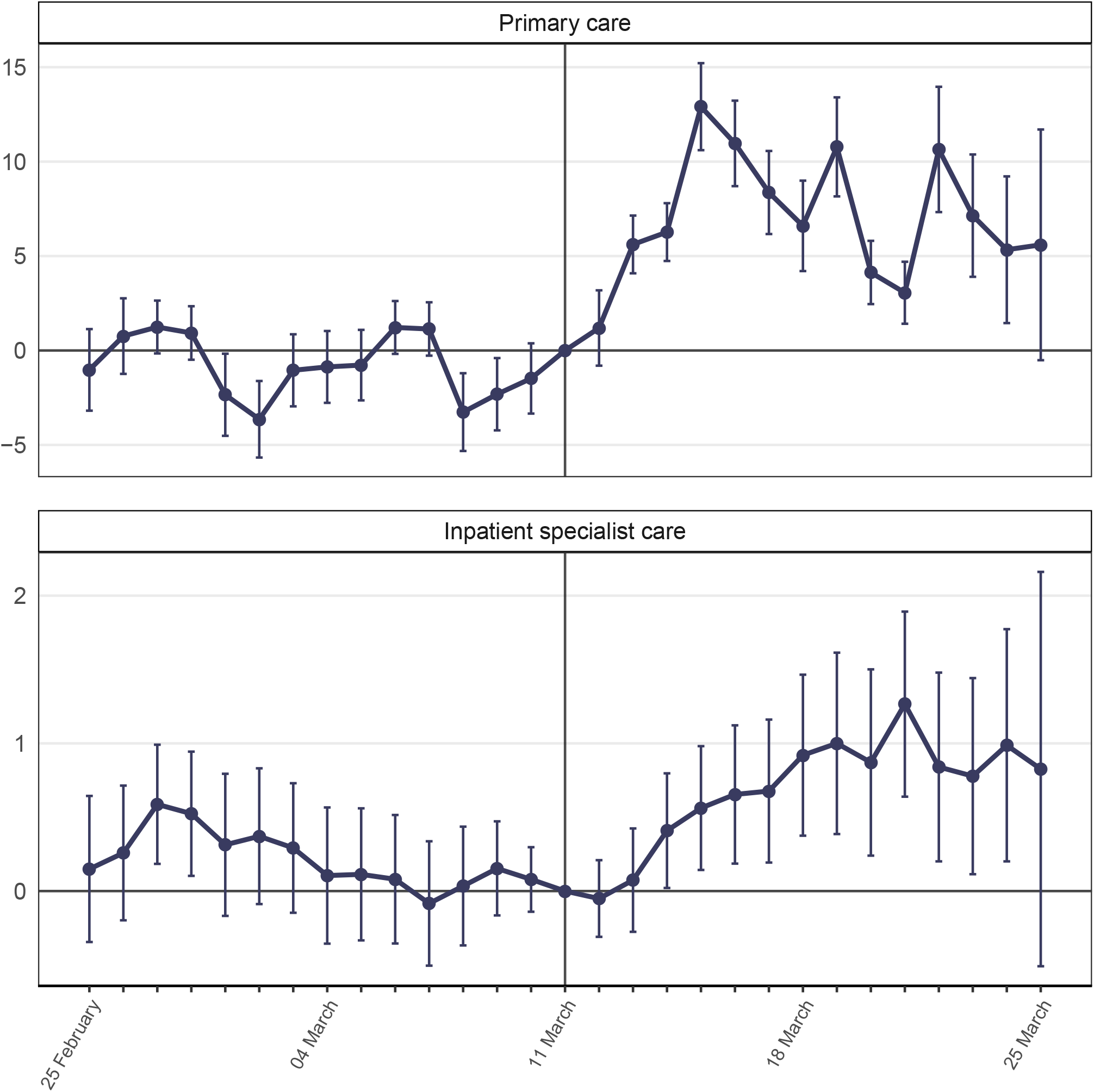
Impact of the information shock on utilization of primary and inpatient care Estimated daily difference in health care use per 1,000 HCWs (95% CI) for those vaccinated with AstraZeneca compared to the matched comparison group, prior to and after the information shock on March 11^th^. Standard errors are clustered on individual.

### Information shock and health care use by age and sex

Health care use, both before and after March 11^th^, was found to differ by age and sex for those vaccinated with AstraZeneca (Fig. 3, Table A1). Prior to March 11^th^, women had higher utilization rates of primary and specialist care than men (Fig. 3). Compared to utilization rates prior to March 11^th^, primary health care use among women aged 18-44 increased with 10.6 per 1,000 (79 %) during the first week after March 11^th^ and with 11.4 per 1,000 (85 %) during the second week (Table A1). For women aged 45-67, the corresponding increase was 6.7 per 1,000 (51 %) in the first week and 6.6 per 1,000 (51 %) in the second week (Table A1).

**Figure 3.**
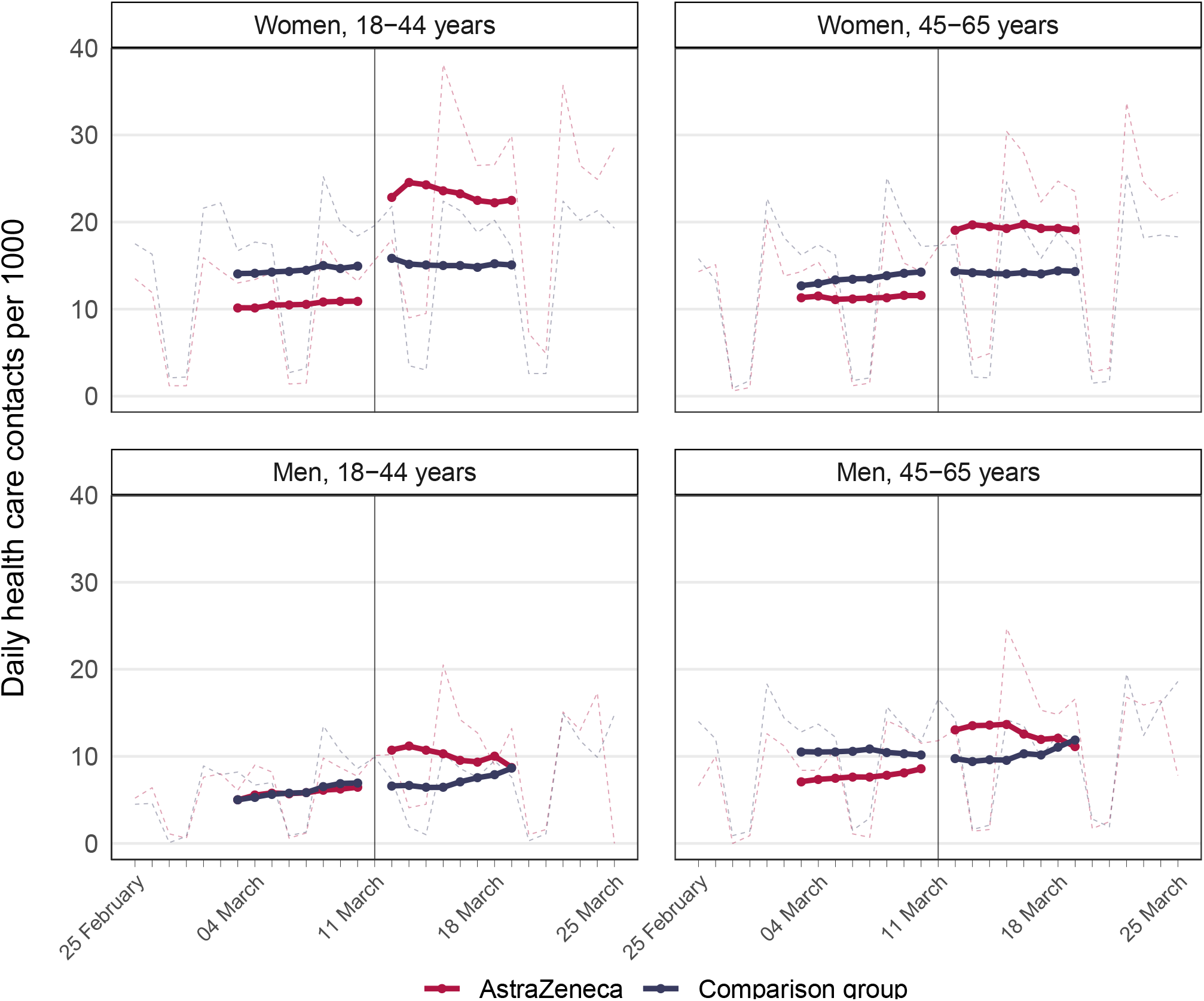
Average health care utilization after the information shock by age and gender Daily (dashed lines) and 7-day rolling average (solid lines) of daily health care use after March 11^th^. The 7-day rolling average was calculated separately before and after March 11^th^ to visualize the change in health care use after the media attention. For days prior to and including March 11^th^, the 7-day rolling average equals the mean of health care use per 1,000 HCWs for the given day and the six preceding days, while it for days after March 11^th^ equals the mean of health care use per 1,000 HCWs for the given day and the six subsequent days.

A similar increase in use of primary care of 4.6 per 1,000 HCWs (75 %) was found for men aged 18-44 the first week after March 11^th^, and 2.1 per 1,000 (34 %) in the second week, compared to the pre-period (Table A-A1). For men aged 45-67, the corresponding increases were 5.7 per 1,000 (59 %) and 2.2 per 1,000 (23 %) (Table A1). However, the overall average use of health care after March 11^th^ was lower among men than women in both age groups (Fig. 3).

### Information shock and health care use by occupation

The impact of the information shock on health care use was also found to differ across HCW occupational groups (Fig. 4, Table A2). While primary health care use for physicians increased with 3 per 1,000 (50 %) in the first week after March 11^th^, the corresponding increase for HCWs working as cleaners was 9.8 per 1,000 (114 %) (Table A2). Health professionals, health associate professionals, and personal care workers had similar increases in primary care the first week after March 11^th^, ranging from 59 % to 65 %. The results for inpatient specialist care were more inconsistent, however the only statistically significant result on a 5 % confidence level in the second week after March 11^th^ was found for personal care workers, with an increase of 1.1 per 1,000 (91 %) (Table A2).

**Figure 4.**
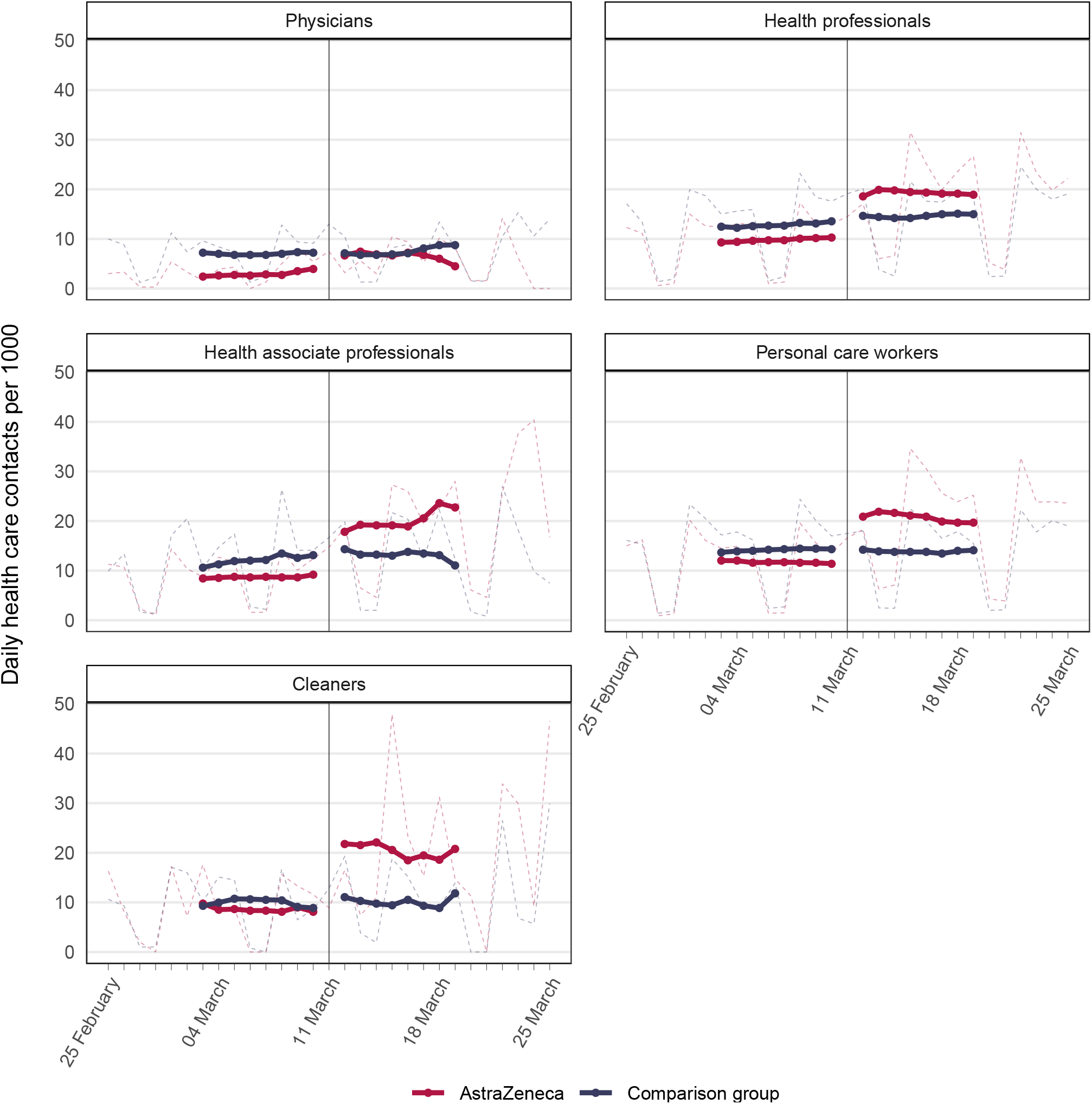
Average health care utilization after the information shock by occupational groups Daily (dashed lines) and 7-day rolling average (solid lines) of daily health care use before and after March 11^th^ for occupations with different work tasks, based on the first digit in ISCO-08 classification (except for physicians). The 7-day rolling average was calculated separately before and after March 11^th^ to visualize the change in health care use after the media attention. For days prior to and including March 11^th^, the 7-day rolling average equals the mean of health care use per 1,000 HCWs the given day and the six preceding days, while it for days after March 11^th^ equals the mean of health care use per 1,000 HCWs the given day and the six subsequent days.

## Discussion

Our study of 99,883 health care workers (HCWs) who had received the AstraZeneca vaccine shows that the daily use of primary care increased by 66 % and inpatient specialist care increased by 20 % during the first week after the information shock of potentially fatal side-effects on March 11^th^, 2021 (Table 2). This corresponded to 8.2 additional primary care consultations per 1,000 vaccinated HCWs each day, and 0.2 additional hospitalizations per 1,000 each day for a week after the information shock (Table 2). The largest increases in primary care consultations the first week after March 11^th^ were found for young women and cleaners working in the health care sector, while older men and physicians were found to be the least affected by the information shock (Table A1, Table A2).

This study both confirms and sheds new and important light on previous studies of the communication of vaccine-related side effects. A randomized experimental study on 292 individuals in Canada found that the individuals who were presented with advantages of influenza vaccination reported fewer systemic side effects than the individuals who were communicated negatively framed vaccine-related side effects.^16^ Similarly, Trogstad et al. ^4^ also found that 5,132 AstraZeneca vaccinated individuals in Norway were more likely to report skin bleeding in surveys than 3,416 individuals vaccinated with mRNA-vaccines, and the authors discuss that the difference may be due to awareness bias after March 11^th^ when the fatal side effects of AstraZeneca vaccination were communicated. In line with previous studies, we have demonstrated an increase in health care use immediately following negatively framed communication of side effects (Table 2). Hence, our findings may indicate that the information shock introduced vaccine-related awareness and that future studies of side effects and health-seeking behavior need to take negatively framed media coverage into account – and this is likely true for any type of vaccine.

To our knowledge, this study is the first to explore changes in utilization of primary and specialist care for vaccinated individuals after such an information shock concerning vaccine-related side effects. For health authorities, our findings imply that both primary and specialist health care services may need to prepare for handling situations of high media attention on possible severe side effects, even though the risk of experiencing the severe side effect is extremely low. The excess thrombolytic events after AstraZeneca vaccination were estimated to be 11 per 100,000 ^17^, and our results thus add to an intriguing literature on humans’ responsiveness to negative events of very low probabilities.^18^ The largest increase in primary care use was found among young women which supports existing theory arguing that women are more risk averse than men.^19^ The second and third largest increase in primary care consultations was found among young men and among older women (Table A1). The perceived risk may have been high for all three groups as the public was informed that a majority of the most serious cases of suspected vaccine-related side effects occurred among young individuals, mostly women.

Furthermore, our results for the first week after March 11^th^ showed a larger and more consistent increase in primary care consultations than hospital admissions (Table 2, Fig. 2), which may be explained by the former acting as a gatekeeper to specialist care. However, the following week, we observed an increase in overnight hospital stays (Table 2, Fig.2), especially among women (Table A1). While this may indicate that a larger share of those vaccinated required specialist care in this period, it may also indicate that primary care physicians’ threshold for referring vaccinated patients to specialist care was lowered due to the information shock, even though the vaccinated physicians’ own health care use was little affected (Fig. 4, Table A2).

The observed increase in health care use is driven, at least partly, by behavioral responses to information shocks, which implies that general utilization data may be more suitable as an indicator of an individual’s overall health rather than somatic health. In these situations, only hard outcomes, eventually mainly death, can be trusted to reflect relevant information about the latent health of the patient. In the current debate about side effects of AstraZeneca, it is thus interesting that a study based on utilization data found an increase in milder diagnoses, but a *decrease* in mortality.^17^ As our results have shown, using these utilization data for real-time monitoring and evaluation of previously unknown vaccine side effects may be biased without carefully considering information shocks and awareness bias on individual health-seeking behavior.

Important strengths of our study include the large sample size, the inclusion of both vaccinated and unvaccinated HCW, as well as the use of a comparison group. Another strength is the use of routinely collected, daily nation-wide data from health registers that are mandated by law. Our methodological approach, where we studied health care use both before and after the information shock on March 11^th^ also add to the strengths of the study, as it allowed us to provide estimates of what may happen to health-seeking behavior in the wake of unexpected news about side-effects of medication, while controlling for time-variant confounders, such as seasonal changes to health care use that impact both groups to the same extent. However, there are also certain limitations to our analysis. First, the vaccine was not administered to HCWs with infection and fever on or before the vaccination day, and we did not have information to exclude HCWs with such symptoms from the comparison group. This may have led to selection of somewhat healthier individuals into our treatment group and may explain why the level of health care use prior to March 11^th^ was slightly higher for the comparison group than for the vaccinated group. However, the difference in health care use per 1,000 persons between the vaccinated HCWs and the HCWs in the comparison group before the information shock on March 11^th^ was small compared to the difference after March 11^th^ (Fig. 1; Fig. 3), which makes it unlikely to have led to considerable bias in our effect estimates. Second, we do not attempt to shed any light on whether the increase in health care use led to better health, and to what cost. To assess this, further research should strive to quantify the health gain and costs of increases in health care utilization under such information shocks. Third, our data only contains registered consultations and hospital admissions. This may have led to an underestimation of the results as our study population consisted solely of HCWs who could potentially have received second opinions from colleagues, particularly in periods of greater pressure on the health care services. In addition, due to our specific study population and context (HCWs in Norway), the external validity of our results may be limited. Accordingly, our stratified analysis showed different impacts of the information shock for different occupations, with occupations not requiring formal health education, such as cleaners, having a larger increase in health care use than other occupations working in the health care sector. This may imply that the individual level of health literacy could have been a contributing factor when responding to the information shock, and hence that our estimates of the impact of the information shock again may be underestimated compared to the impact in the general population.

## Conclusions

By studying HCWs vaccinated with the AstraZeneca vaccine, we have shown that the unexpected reports of fatal vaccine-related side effects on March 11^th^, 2021, led to a significant increase in utilization of primary and specialist care. Our study illustrates that health services should be prepared for increases in consultations in the wake of future information shocks about negative events with low probabilities, like possible vaccine-related side effects.

## Data Availability

The individual-level data used in this study is not publicly available due to privacy laws.

**Uncategorized References**

## List of abbreviations

EMA: European Medicines Agency
HCW: Health care worker
NIPH: Norwegian Institute of Public Health

## Notes

### Competing Interest Statement

The authors have declared no competing interest.

### Funding Statement

The study was funded by the Norwegian Institute of Public Health. No external funding was received.

### Summary of Updates

Study sample (treatment and comparison groups) redefined and all statistical analyses reconducted with updated sample; sections on methods and results updated to reflect revisions; all figures and tables updated; Appendix updated.

