## Appendix for "Utilization of health care services before and after media attention about fatal side effects of the AstraZeneca vaccine: a nation-wide register-based event study"

**Figure A1:** Full sample size and exclusion criteria.

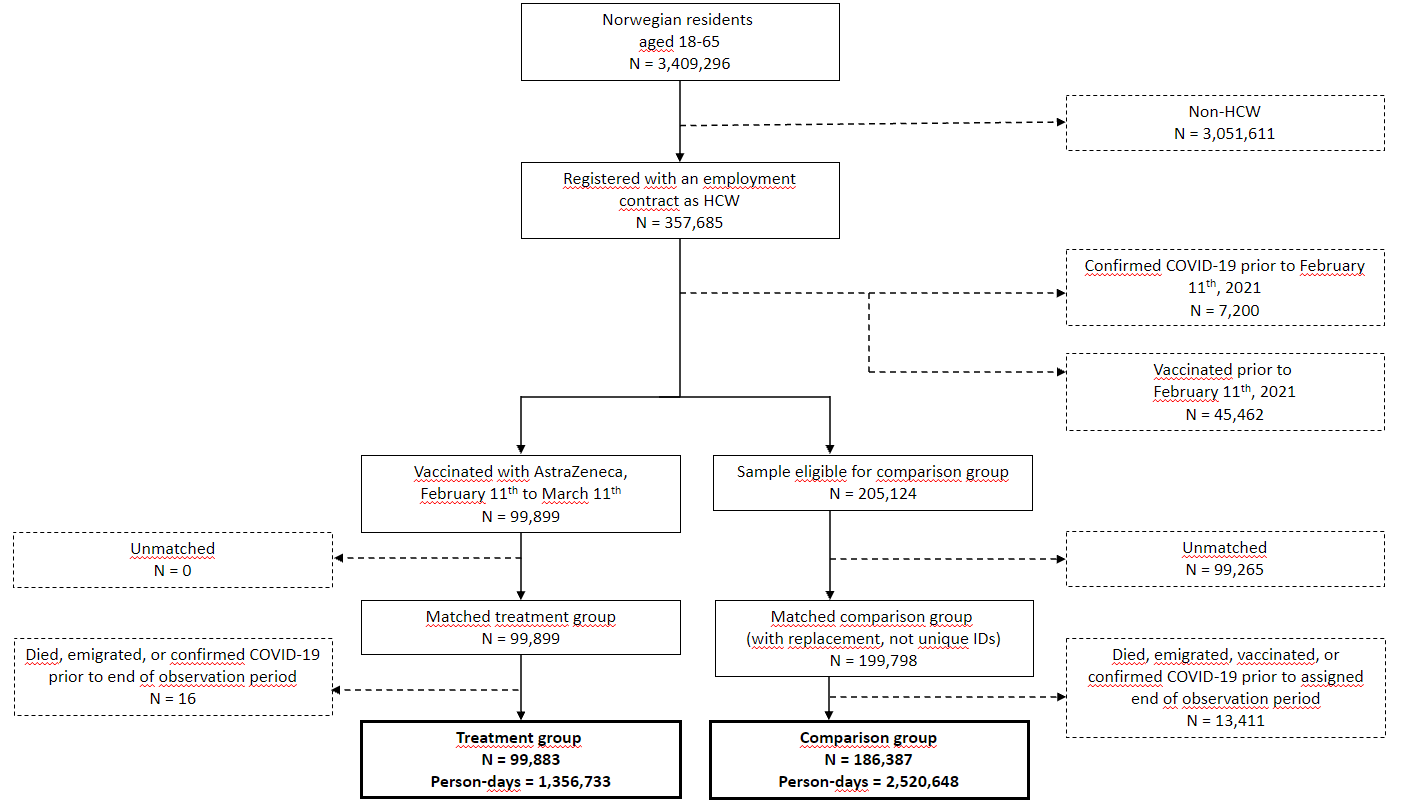

Figure note: Due to the matching procedure with replacement, there is a discrepancy in the sample size of the HCWs eligible for matching and of the final matched comparison group, where the former consists of unique individuals while the latter does not.

***Details on propensity score matching***

To mimic a situation with quasi-experimental as-if random distribution of vaccines we constructed an unvaccinated comparison group using the propensity score matching method. We matched each health care worker (HCW) vaccinated with AstraZeneca to two unvaccinated (as of February 11^th^, 2021) HCWs. The propensity scores were created via a logit model that estimated the likelihood of receiving the AstraZeneca vaccine, imitating a randomized distribution of vaccines, using the following covariates: age (treated as a categorical variable); sex; the county in which the main employment was registered; occupation categories; and industrial categories (see appendix of Molvik et al. for exact definitions). We also controlled for utilization of primary (consultations and outpatient hospital contacts) and specialist (inpatient hospital contact) health care in November and December 2020, defined as number of weeks with at least one contact. This allowed us to balance the pre-trend in the control and treatment groups, without using post-treatment information.

Following convention, we used the nearest-neighbor (NN) method to balance the treatment and comparison groups.^20^ The algorithm minimizes the absolute difference between the propensity scores of the vaccinated individual and its two controls. To further increase the quality of our matches, we matched using replacement. This implies that individuals in the target set could be matched to treated individuals more than one time. Duplicates in the comparison group means that the data is no longer independent. We adjusted for the dependence in our data using clustered standard errors in the main analysis.

***Supplementary tables***

**Table A1:** Impact of the information shock by age and sex

|  |  | **Primary care** | | | |  |  | **Inpatient specialist care** | | | | |  | |
| --- | --- | --- | --- | --- | --- | --- | --- | --- | --- | --- | --- | --- | --- | --- |
|  | Period after March 11^th^ | β | | St. err | % Relative diff (β) | Relative diff (St. err.) |  | | β | St. err | % Relative diff (β) | | | Relative diff (St. err.) |
| **Women** |  |  | |  |  |  |  |  | |  | |  |  | |
| Age 18-44 | First week | 10.6*** | 0.55 | | 79 | 4.1 |  | 0.4 | | 0.23 | | 22 | 14.5 | |
|  | Second week | 11.4*** | 0.84 | | 85 | 6.2 |  | 0.8** | | 0.34 | | 51 | 21.2 | |
| Age 45-65 | First week | 6.7*** | 0.58 | | 51 | 4.4 |  | 0.2 | | 0.26 | | 21 | 29.1 | |
|  | Second week | 6.6*** | 0.88 | | 51 | 6.7 |  | 0.9** | | 0.35 | | 100 | 40.2 | |
| **Men** |  |  |  | |  |  |  |  | |  | |  |  | |
| Age 18-44 | First week | 4.6*** | 0.78 | | 75 | 12.9 |  | 0 | | 0.17 | | 6 | 56.6 | |
|  | Second week | 2.1* | 1.22 | | 34 | 20 |  | 0.2 | | 0.24 | | 53 | 82.3 | |
| Age 45-65 | First week | 5.7*** | 1.15 | | 59 | 12.1 |  | 0.1 | | 0.6 | | 13 | 52.4 | |
|  | Second week | 2.2 | 1.7 | | 23 | 17.9 |  | 1 | | 0.98 | | 90 | 86.1 | |

*Notes:* Differences-in-differences estimates for the change in daily health care use per 1,000 HCWs for different health care services before and after March 11^th^ for HCWs vaccinated the past 14 days. Standard errors (St. err.) are clustered on individuals. The pre-period (health care utilization after vaccination the 2 weeks prior to March 11^th^) is reference period in all regressions. In addition to the presentation of results as absolute differences per 1,000 HCWs we also presented relative differences (i.e., in percent) by dividing the absolute estimate (and corresponding standard error) for each of the post periods by the health care use rate of the comparison group in the pre period (and multiplying with 100). Significance levels: * <0.1; ** <0.05; *** <0.01)

**Table A2:** Impact of information shock on health care use after vaccination by occupational group

|  |  | | **Primary care** | | | |  |  | **Inpatient specialist care** | | |  |
| --- | --- | --- | --- | --- | --- | --- | --- | --- | --- | --- | --- | --- |
|  | Period after March 11^th^ | | β | St. err | % Relative diff (β) | | Relative diff (St. err.) |  | β | St. err | % Relative diff (β) | Relative diff (St. err.) |
| **Physicians** | First week | | 3*** | 1.05 | 50 | | 17.4 |  | 0.9 | 0.56 | 66 | 42.2 |
|  | Second week | | 2.3 | 1.63 | 38 | | 27.1 |  | 0.1 | 1 | 11 | 75.5 |
| **Health professionals** | First week | | 7.2*** | 0.65 | 59 | | 5.3 |  | -0.2 | 0.25 | -13 | 21.1 |
|  | Second week | | 8.1*** | 1.07 | 66 | | 8.8 |  | 0.1 | 0.41 | 9 | 34.9 |
| **Health associate professionals** | First week | | 7.2*** | 1.66 | 64 | | 14.7 |  | -0.4 | 0.64 | -52 | 73.8 |
|  | Second week | | 13.8*** | 2.96 | 122 | | 26.2 |  | -0.3 | 0.52 | -30 | 60.4 |
| **Personal care workers** | First week | | 8.8*** | 0.47 | 65 | | 3.5 |  | 0.4* | 0.21 | 34 | 17.6 |
|  | Second week | | 7.9*** | 0.67 | 58 | | 5.0 |  | 1.1*** | 0.29 | 91 | 24.3 |
| **Cleaners** | First week | | 9.8*** | 3.26 | 114 | | 37.8 |  | 0.3 | 1.10 | 49 | 181.5 |
|  | Second week | | 5.3 | 4.6 | 62 | | 53.3 |  | 3.3 | 2.03 | 544 | 336.1 |

*Notes:* Differences-in-differences estimates for the change in daily health care use per 1,000 HCWs for different health care services before and after March 11^th^ for HCWs vaccinated the past 14 days. Standard errors (St. err.) are clustered on individuals. The pre-period (health care utilization after vaccination the 2 weeks prior to March 11^th^) is reference period in all regressions. In addition to the presentation of results as absolute differences per 1,000 HCWs, we also presented relative differences (i.e., in percent) by dividing the absolute estimate (and corresponding standard error) for each of the post periods by the health care use rate of the comparison group in the pre period (and multiplying with 100). Significance levels: * <0.1; ** <0.05; *** <0.01)
